# Spatial analysis of methicillin-resistant *Staphylococcus aureus* carriage (MRSA) at hospital admission in a livestock dense region

**DOI:** 10.1101/2023.05.01.23289266

**Authors:** Vera H. Arntzen, Sabiena G. Feenstra, Elisa Benincà, T.T.N. Le, Ellen M. Mascini, Marrigje H. Nabuurs-Franssen, Andreas Voss, Agi M. Marik, Eefje de Jong, Welmoed Silvis, Maarten J. Schijffelen, Peter Schneeberger, Joost Hopman, Hester Korthals Altes, Heiman F.L. Wertheim

## Abstract

**Title:** Spatial analysis of methicillin-resistant *Staphylococcus aureus* carriage (MRSA) screening yield at hospital admission in a livestock-dense region

**Purpose:** In the Netherlands, patients with a risk factor for methicillin-resistant *Staphylococcus aureus* (MRSA) carriage, such as foreign hospital stay and contact with livestock (pigs, veal calves and/or broilers) are actively screened upon hospital admission. This study aimed to give insight in the geographical clustering patterns of MRSA carriage among these patients in a livestock-dense region.

**Methods:** A retrospective study was performed using medical records and laboratory results of MRSA screened patients admitted to seven hospitals in the provinces of Gelderland and Noord-Brabant, covering the period from 01/2011 to 02/2017. SaTScan spatial scanning identified cluster areas with an increased MRSA carriage risk in postal codes compared to the surrounding areas.

**Results:** 15 546 patients were included, among which 10.0% (n=1499) were MRSA carriers. Four significant, typically highly pig-dense MRSA carriage hotspots were identified, where the relative risk of carriage ranged from 2.1 to 3.4 compared to the surrounding area.

**Conclusion:** MRSA carriage risk clustered in certain areas, suggesting an association between livestock density (mainly pigs) and the MRSA carriage risk for the screened population at hospital admission. It needs to be explored when proximity (not contact) to livestock should be considered a risk factor. Considering analytical difficulties we encountered it is recommended to harmonize culture methods and data acquisition across hospitals to facilitate analysis for improving MRSA screening policy.

**Impacts:**

- The fraction MRSA carriers among patients with livestock contact notified in the hospital system in this study was 22.7%, showing the effectiveness of the Search and Destroy policy.
- MRSA carriage in the Dutch Province Gelderland is clustered in certain hotspots, located in pig-dense areas.
- Culture methods and data acquisition should be harmonized across hospitals to facilitate analysis for improving MRSA screening policy.

## Introduction

In 2015, 1,6% of invasive *Staphylococcus aureus* isolates from hospitals in the Netherlands were methicillin-resistant, compared to 12,3% and 11,2% in neighboring countries Belgium and Germany (Workgroup Infection Prevention (WIP), 2012). Healthy individuals carrying MRSA form a hazard for the hospital population, which is more vulnerable for mild to severe MRSA infection. The Netherlands has an active search-and-destroy policy for MRSA, described in national guidelines for Dutch hospitals (Workgroup Infection Prevention (WIP), 2012). Currently, asylum seekers, as well as patients who recently stayed in a foreign hospital, had contact with pigs, veal calves or broilers, or have/are children adopted from abroad, undergo screening for MRSA carriage upon hospital admission. Swabs collected from nose, throat and perineum are subjected to microbiological testing. Until these are found negative, strict isolation is put in place: the patient has an individual room and hospital staff wear personal protective equipment during contact with the patient.

Following new insights in MRSA carriage risk factors, the search and destroy policy evolved since its start in the 1970s, when the main focus was on persons that previously stayed in a foreign hospital. From 2001 to 2006, the annual MRSA incidence increased more than three-fold, due to the emergence of livestock associated (LA) MRSA (van Rijen et al., 2009). Initially, it was found in Dutch pigs and persons having contact with pigs (Voss et al., 2005), and later in other reservoirs. Exposure to occupationally farmed pigs and veal calves is a risk factor leading to screening at hospital admission since July 2006, and exposure to broilers since October 2012. Although transmission to humans has also been reported for horses in equine clinics, turkey, goats and sporadically for feedlot and dairy cattle, these are not included in the current risk factors for screening (Loncaric et al., 2013; Tokateloff et al., 2009; Van den Eede et al., 2013; van Duijkeren et al., 2016; van Duijkeren et al., 2010; Weese et al., 2012). According to the revised guideline of 2012, outpatients are not screened anymore. This policy effectively controls MRSA-spread in Dutch hospitals (van Rijen et al., 2009), and is cost-effective (van Rijen & Kluytmans, 2009). Although the screening program as a whole is effective, little is known about the relative contributions of livestock types to the risk of MRSA carriage.

Active screening is cumbersome for both patients and hospitals, especially in livestock-dense regions. Insight in the spatial/geographical aspects of MRSA positivity at hospital admission might help mapping risk patterns. A major livestock-dense region is located in the East of the Netherlands and covers the complete province of Gelderland and the adjacent northern part of the Province of Noord-Brabant. This explorative study aims to give hospitals and local public health institutes insight in the collective screening results of hospitals in the region and to describe the geographical differences in the MRSA carriage risk among patients screened for MRSA at hospital admission. In this paper we address the following questions:

1. How many patients with a MRSA carriage risk factor, screened at hospital admission over the period 01/2011-02/2017 in a livestock-dense area in the Netherlands, are MRSA carrier?
2. Are there MRSA carriage hotspots within the livestock-dense area in the Netherlands, where the MRSA carriage risk among screened patients is significantly higher than in the rest of the included area?
  a. Where are these hotspots located?
  b. What densities of pigs, broilers and/or veal calves characterize these hotspots?

## Methods

### 1. Design

A retrospective study was performed using medical records and laboratory results from patients admitted to six Gelderland hospitals and one hospital in the province Noord-Brabant (Figure 1). Three hospitals in the region were not included: for two this was due to issues in extracting data from their laboratory systems (Appendix 1), and one was unable to participate. Within the province Gelderland, the eventually included hospitals cover 50.9% of all hospital beds in the province (n=2631) (Hastie, 2009).

**Figure 1.**
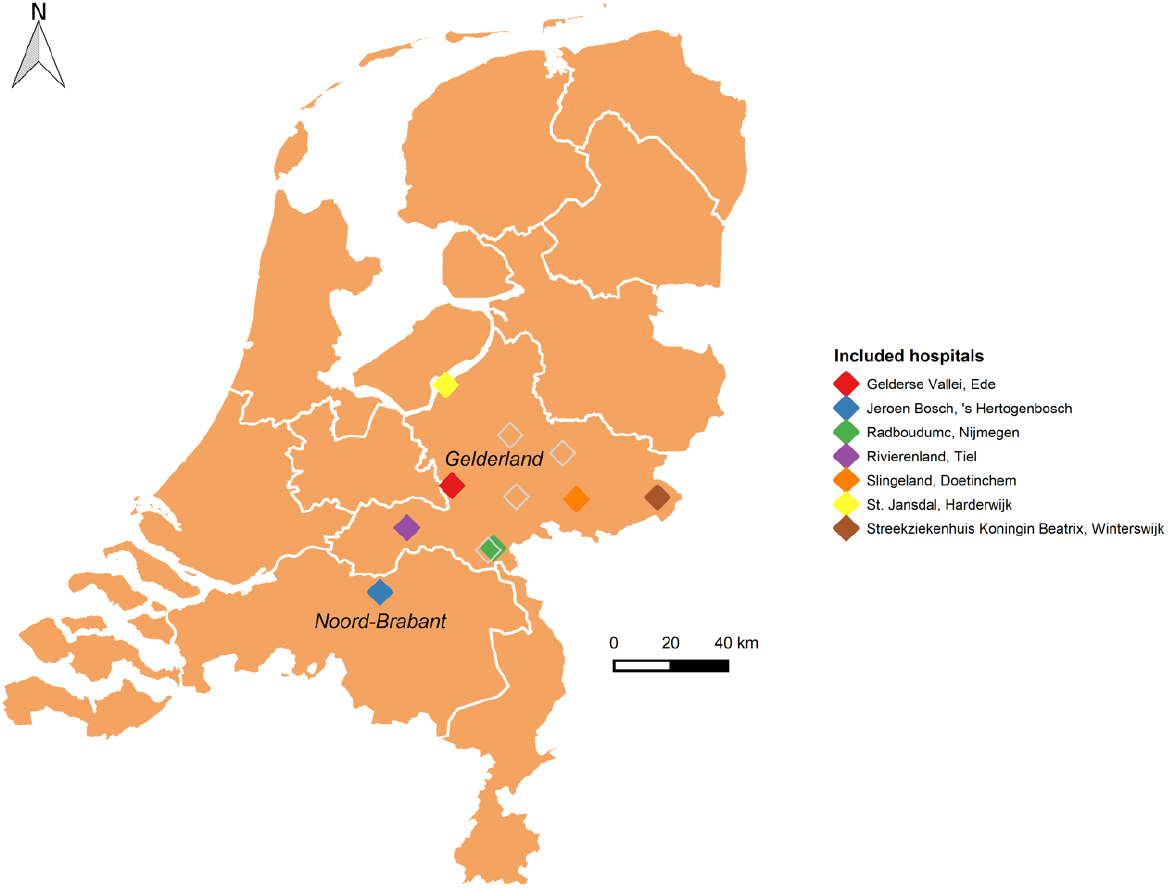
Hospitals in the Netherlands included in this study. Three hospitals in the Province of Gelderland (black border line) were not included in the analysis (open diamond boxes), of which one has two locations; one hospital in the northern part of the Province of Noord-Brabant was included as well.

The study population consisted of all patients with a MRSA carriage risk factor who were screened for MRSA carriage at outpatient visit or admission to one of the included hospitals during the six year-period 01/2011 – 02/2017. The screening policy applies to those having contact with broilers, pigs or veal calves, foreign hospital stay, asylum seekers and adopted children. The outcome of interest was the risk of MRSA carriage among screened patients, i.e., the proportion of screened patients that were found to be MRSA carrier within the study period. A patient was defined as a unique patient per hospital, that could either be MRSA carrier when there was at least one MRSA-positive test result for which material was taken during the study period, or a non-carrier, when there were only MRSA-negative results during the study period. Independent variables were pig, broiler and/or veal calve density in a four-digits postal code.

### 2. Ethical considerations

All locally required research committees agreed with this study (METC dossier 2017-3319). Informed consent of patients was not needed. Patients that previously raised objections against the use of data for scientific studies, were excluded. Unique patient numbers were recoded, age was analysed in 5-years categories and postal codes were only provided with the first four digits for sufficient anonymity.

### 3. Data collection

Data on MRSA carriage were collected from the medical microbiology laboratories of hospitals. This included available patient information as well as test-related information of all laboratory tests for MRSA, both screening and coincidental findings of MRSA, of patient’s material collected in the period 01/2011-02/2017. Laboratory tests for MRSA are based on detection of the species S. aureus and the mec-A-gene coding for the modified penicillin-binding protein PBP-2a, which decreases affinity for antibiotics such as methicillin. Swabs from throat and nose were either combined in one culture or cultured separately, and tested once or twice, yielding different numbers of tests per screening episode. Thus, per screening moment, usually two to six to cultures were tested. Each test corresponded to one row in the original data. Patient information included a unique identifier (ID) per patient per hospital, the four-digit postal codes of the home address, sex, age group (per five years) and patient or employee status. Test-related information included the date on which patient material was obtained, hospital admission date, the patient material type (nose/perineum/throat-swab or else), the procedure (screening or coincidentally found MRSA in clinical cultures), and type of request (for example by a General Practitioner or a nursing care home). The hospital admission date was used to distinguish hospital admission from outpatient visit. When part of screening, additional information included possible MRSA carriage risk factors.

Data were not uniformly available at all hospitals. With data managers and/or medical doctors, the appropriate variables were selected to derive the relevant information. While most variables of interest were conveniently stored in columns, MRSA carriage risk factors, contact tracing and types of MRSA-strains often either needed to be hand-coded from open-text clinical data or were lacking. A coding scheme was set up per hospital and verified by the data manager and/or medical doctor of the specific hospital (data not shown).

Population statistics of 2015 and total population counts per age/sex group of 2011-2016 were available per four-digit postal code through Statistics Netherlands (Statistics Netherlands, 2016a, 2016b). Farm locations and corresponding counts per livestock type (broilers, veal calves, pigs) were available from the Dutch Agricultural Census Register (Dutch Ministry of Agriculture and Environment, 2015) and aggregated to postal code-specific totals per livestock type. A list of livestock-associated spa- and MLVA-types was available through RIVM.

### 4. Inclusion criteria

All screening tests for MRSA performed by the hospital laboratories on patient material collected in the period 01/2011 up until 02/2017, as well as coincidental findings in routine bacterial cultures on patient materials from that period, were included (Figure 2). Further inclusion criteria were: a conclusive test result, availability of a four-digits postal code in the Netherlands as well as presence of a unique ID. Tests performed for contact tracing were excluded, as they would not relate to exposure in the patients’ living area, but to exposure elsewhere - as in the hospital itself for example. When multiple MRSA positive tests occurred for a single patient, only the first positive was considered as an event, because this may concern a single colonization in a person or repeated exposure to the same environment. The data allowed to define unique patients per hospital only, since patient IDs differ between hospitals and data were anonymous. From every unique patient per hospital, the first MRSA-positive test result in the study period was used, or when this was lacking, the first negative test. Patients for whom the first MRSA-positive test was a coincidental finding, were beyond the scope of this study since these patients did not have a MRSA carriage risk factor included in the search-and-destroy policy. Patients screened for MRSA upon hospital admission were included for analysis, and referred to as screened patients.

**Figure 2.**
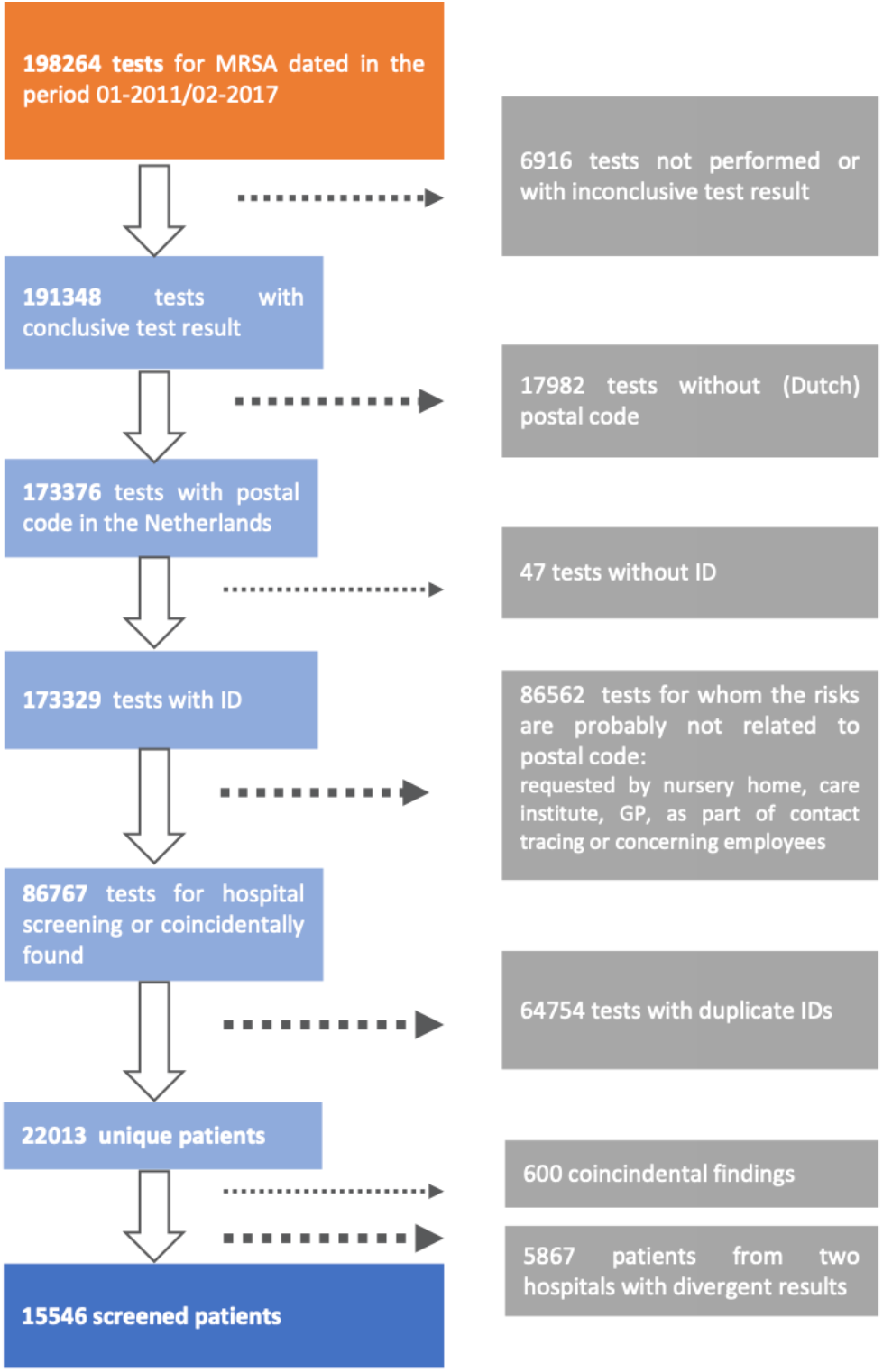
Inclusion criteria of patients screened for MRSA carriage.

### 5. Data analysis

A pilot study was performed with data available from the department of Medical Microbiology at the Radboudumc, followed by main analyses with data covering the Province Gelderland and the North of the provinces Noord-Brabant and Limburg. Seasonality of MRSA carriage (Hassoun et al., 2015; Leekha et al., 2012; Mermel et al., 2011) was not considered in our data since we did not reveal any time trends. All analyses were performed in Statistical Software Program R, version 3.6.1,and RStudio, version 1.0.153 (R Development Core Team, 2016; RStudio Team, 2016).

#### 5.1 Descriptive analyses

Patient characteristics and the fraction MRSA carriers were summarized. The age/sex distribution was shown in a figure using 5-year age groups (Appendix 2). For practical reasons, in the following, a division into only six age groups (Table I) was considered. Screened patients and screening intensity (e.g. MRSA screened patients per 100 000 inhabitants) were mapped per four-digits postal code (Figure 3). Since data on hospital catchment was not generally available, the population count was chosen as denominator.

**Table I.**
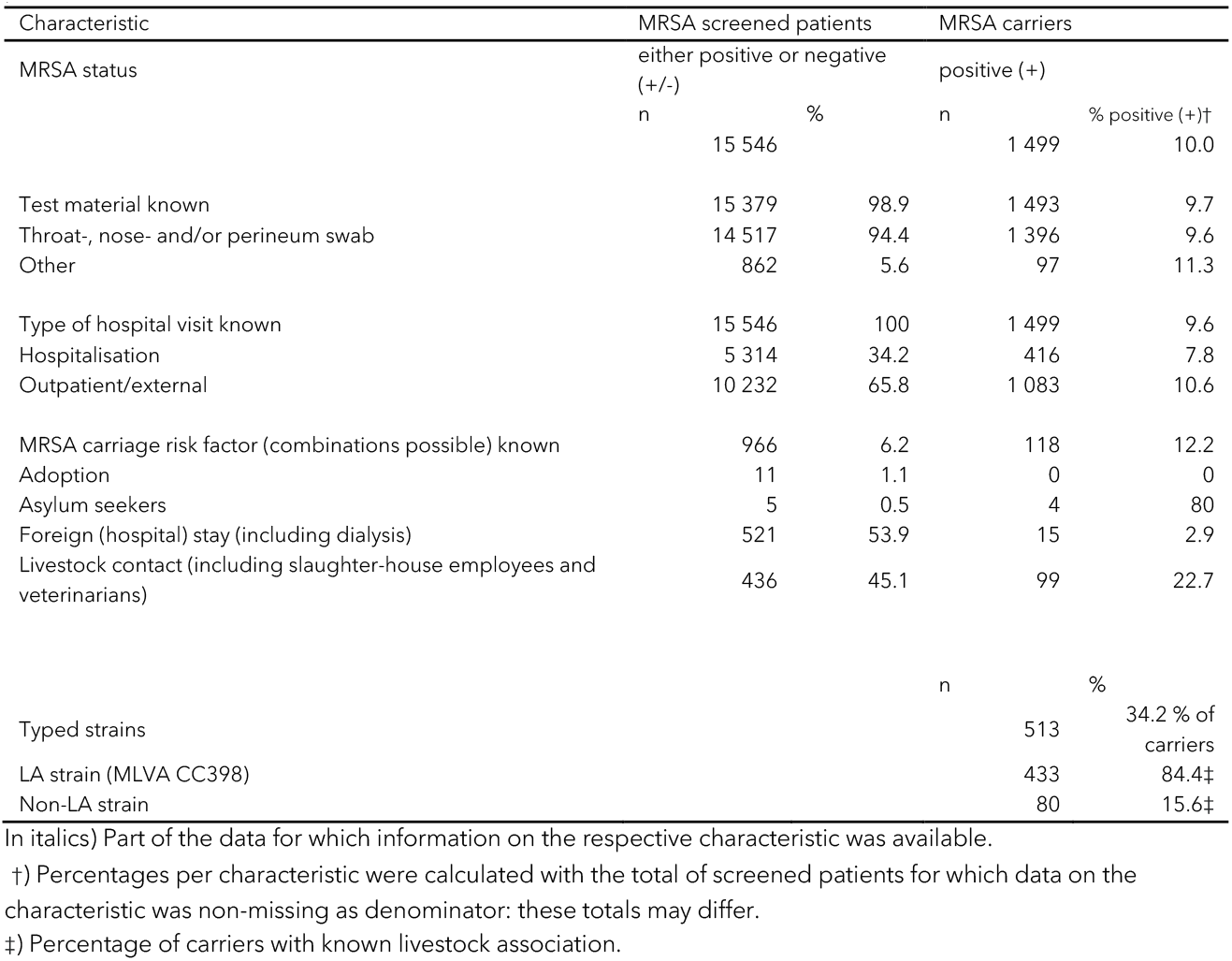
Characteristics of MRSA screened patients included in this study. Not all variables were available for all patients.

**Figure 3.**
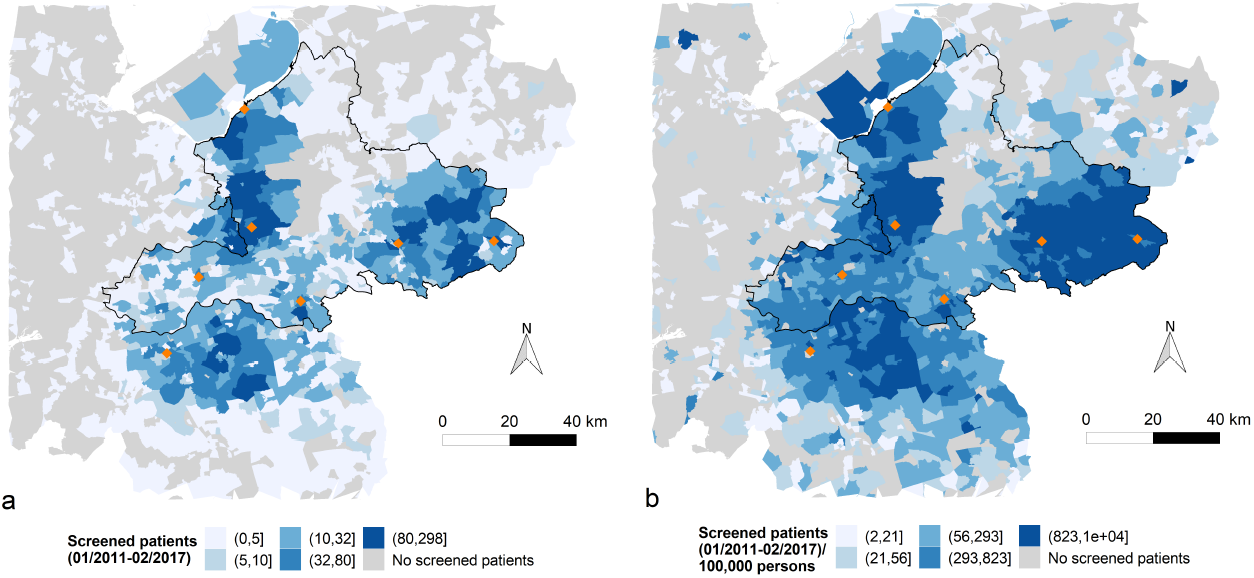
MRSA screening per postal code in the period 01/2011-02/2017. (a) Absolute number of patients screened per four-digits postal code. Categories are the 6-quantiles (areas with count 0 excluded) where the smallest categories were aggregated to one level ([1,5)) for anonymity. SaTScan analysis was performed for four-digits postal codes with at least 10 screened patients only; (b) relative number of screened patients, e.g. screened patients per 100 000 inhabitants of a four-digits postal code (2015). Categories are the 5-quantiles (areas with count 0 excluded). Orange (filled) diamonds represent (included) hospitals. The border of the Province Gelderland is indicated in black.

#### 5.2 Spatial analysis

To study spatial differences in the MRSA carriage risk at hospital admission per four-digits postal code, the adjusted MRSA carriage incidence among screened patients was mapped, for four-digits postal codes. To guarantee anonymity of test results and to prevent unreliable estimates when the denominator is small, incidences are shown only for postal codes with at least 10 screened patients (Figure 4a). MRSA carriage incidence was calculated as the cumulative number of MRSA carriers divided by the cumulative number of screened patients over the study period in a four-digits postal code, times 1000.

**Figure 4.**
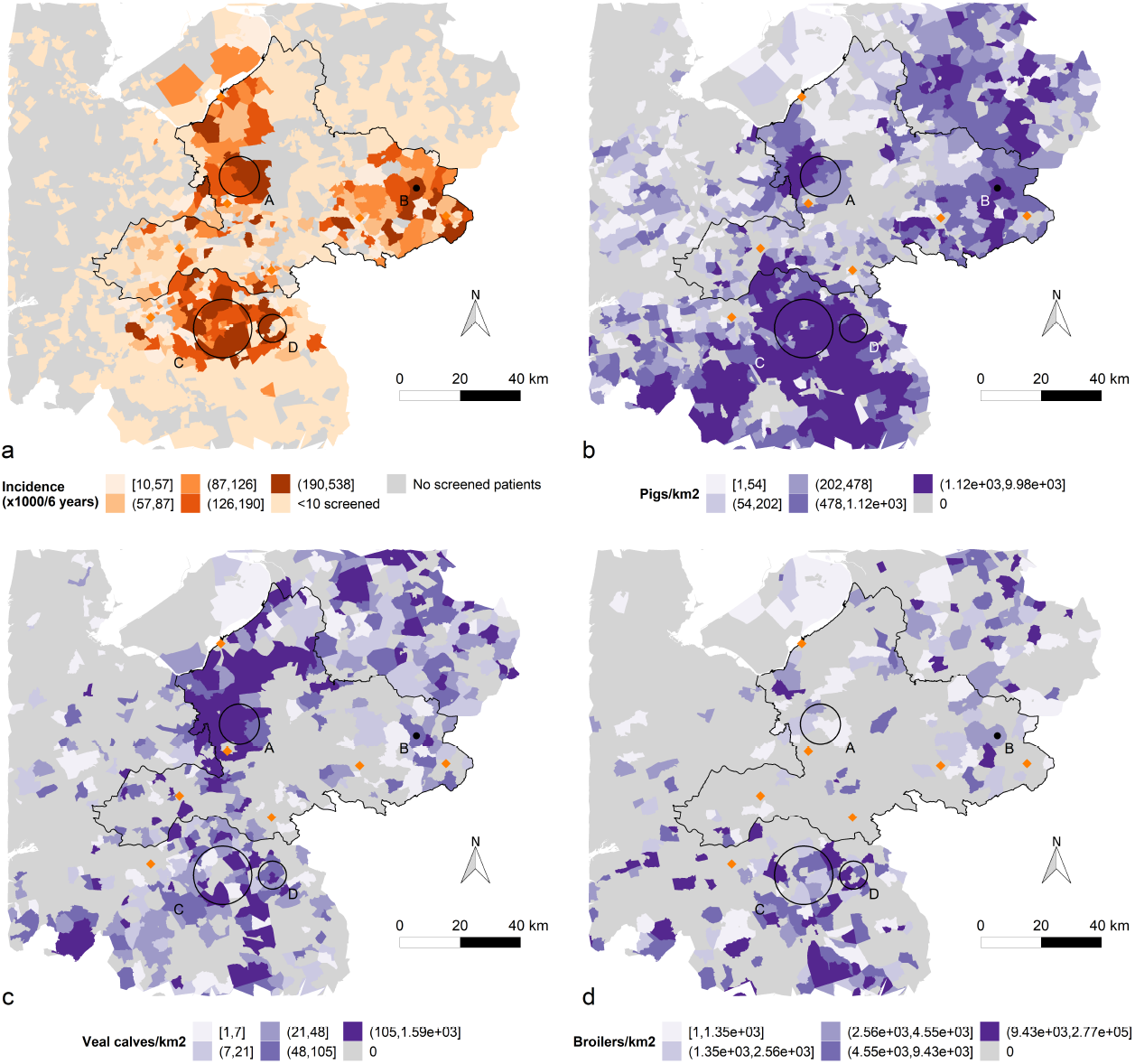
MRSA carriage and livestock density. MRSA carriage hotspots identified by SaTScan Poisson cluster analysis aresuperimposed on maps with (a) adjusted MRSA carriage incidence, e.g. MRSA carriers per 1 000 screened patients in a four-digits postal code in the period 01/2011-02/2017; (b) pigs/km^2^ per four-digits postal code; (c) veal calves /km^2^ per four-digits postal code; (d) broilers/ km^2^ per four-digits postal code. In all maps, categories are the 5-quantiles (areas with count 0 excluded). In these MRSA carriage hotspots, indicated as black circles, the MRSA carriage among screened patients was respectively 1.9 to 4.4 times higher than in the other included area, adjusted for age and sex (Table II). Orange or blue (filled) diamonds represent (included) hospitals. The border of the Province Gelderland is indicated in black.

Spatial patterns of MRSA carriage were analyzed with clustering analysis software program SaTScan (Kulldorff, 2015), run from R with the package ‘rsatscan’ (Kleinman, 2016). SaTScan iteratively centered a flexible window with varying radius in the centroid of a postal code, and compared counts of observed and expected cases inside and outside the window. We allowed clusters of at most 10 km radius, containing at most 10% of the population at risk. This maximum radius was set, in order to limit the heterogeneity in estimated MRSA carriage risk within a cluster. Here, events were MRSA carriage, and the population consisted of the screened patients. The expected case count was calculated by assuming a Poisson distribution (discrete Poisson model (Kulldorff, 1997; Kulldorff, 2015). For details about the scanning procedure, see Appendix 2. Densities of the livestock types associated with MRSA risk (pigs, veal calves and/or broilers) were transformed to a log10 scale for interpretability of the maps.

## Results

### 1. Inclusion of patients

In total, 198 264 MRSA test results were obtained from nine hospitals in the Netherlands, of which 86 767 tests met the inclusion criteria (Figure 2). From these, one test was selected per patient per hospital, namely the first positive or, if the patient never tested positive, the first negative test. This yielded 22 013 patients that were unique per hospital. For two hospitals (A and I, Appendix 3) the number of tests and/or the percentage MRSA positive patients deviated largely from the other hospitals, probably due to differences in data monitoring practice. We excluded the data from these hospitals, such that in total, 15 546 patients from seven hospitals were included (Figure 1). These patients either had a MRSA carrier or non-carrier status.

### 2. Descriptive analyses

Most screened patients were in the categories 20-64 years old (n=6 930, 59.2%), and slightly more females were screened (n=6 103, 52.2%) than males. The age distribution of screened women was multimodal, with peaks in the age groups 20-34 and 45-59 (Appendix 2A).

The reason for screening, i.e. contact with certain livestock and/or foreign hospital or adoption, was rarely reported (n=966, 6.2%) and varied between 0.0 and 53.2% according to hospital (Appendix 3). The main reported MRSA carriage risk factor was hospitalization in a foreign country (n=521, 53.9%), followed by livestock contact - reported for 436 screened patients (45.1%). However, this is based on little data and cannot be generalized for the study population. Figure 3a shows the absolute number of patients per postal code of residence. The screening intensity, i.e. the number of screened patients per 100 000 inhabitants of a four-digits postal code, varied across the study area; it was highest in the north-western and eastern parts of Gelderland (Veluwe and Achterhoek, respectively) as well as the north of Noord-Brabant (Figure 3b). The screening intensity depends on the population count per postal code: with the same absolute number of screened individuals, it is higher in rural than in urban areas. Test material was most often a culture from a throat, nose and/or perineum swab (n=14, 517, 94.4%), the common sampling sites for MRSA screening. It confirms that most patients were indeed part of the screening program (Table I). The majority of specimens was obtained during outpatient visits (n=14 517, 94.4%). Data on type of livestock contact (broiler, pig, veal calve) were not notified systematically. The proportion of MRSA carriers among screened patients with a reported MRSA carriage risk factor was the highest for livestock contact: 22.7% of patients that had contact with broilers, pigs or veal calves were MRSA carrier (n=99).

The fraction of MRSA carriers among screened patients was 10.0% (n=1499, Table I). For 34.2% of these (n=513), the type of MRSA-strain was reported, which was most often a livestock-associated (LA) strain (MLVA CC398 or an associated Spa type, n=433, 84.4%). For other MRSA carriers, the MRSA-strain was either determined but not reported in the hospital/laboratory system, or completely unknown.

### 3. Spatial analysis

We built a four-digits postal code map of the MRSA carriage incidence in the studied area (Figure 4a). The highest incidence was 190 per 1 000 screened patients in the study period. Four significant (P<.05) clusters were detected with a spatial scan using the SaTScan Poisson model. The clusters together contain 13.2% (n = 2050) of screened individuals, yielding 28.0% (n = 419) of the cases.

Centroids of the clusters A and B lay in the province Gelderland; centroids of clusters C and D in the province Noord-Brabant. Clusters mostly covered rural areas, and spatial cluster size ranged from a single four-digit postal code (cluster B) up to almost 10 km (cluster C) radius, the maximum allowed by the scanning method (Table II). Pig, veal calve and broiler densities differed between the clusters and are color-coded based on the quantiles of their distribution (Figure 4b-d). The clustering pattern roughly followed the distribution of pigs in the included area of provinces Gelderland and Noord-Brabant, but not all pig-dense areas included in the catchment areas of the hospitals were covered by clusters. While pigs were densely farmed in clusters B-D, veal calves were densely farmed in clusters A. However, while the pig dense areas are largely covered by the clusters, there are veal calve dense areas outside the identified hotspots (Figure 4b-c, Table II). High broiler density areas were sparser, with a spotty pattern in the provinces Gelderland and Noord-Brabant, with only some broiler-dense four-digits postal codes contained in all clusters (Figure 4d, Table II).

**Table II.** Characteristics of SaTScan Poisson clusters where the MRSA carriage risk is higher than expected by chance

| Cluster | Radius (km) | P | MRSA carriers | Expected | Screened | Relative risk | Pigs | Veal calves | Broilers |
| --- | --- | --- | --- | --- | --- | --- | --- | --- | --- |
| A | 6.7 | P<.001 | 134 | 66 | 686 | 2.1 | +/- | + | - |
| B | 0.0 | P<.001 | 37 | 14 | 142 | 2.7 | + | +/- | +/- |
| C | 9.7 | P<.001 | 228 | 112 | 1 160 | 2.2 | + | +/- | +/- |
| D | 4.6 | .02 | 20 | 6 | 62 | 3.4 | + | +/- | +/- |

Clusters were selected with criterium $P < .05$ . The presence of pigs, veal calves and broilers farming was indicated as follows: '+', livestock-dense; '+/-': some livestock-dense four-digit postal codes inside the cluster; '-': not livestock-dense.

## Discussion

The aim of this study was to map the MRSA carriage risk among those screened upon hospital admission and examine geographical clustering patterns. We found that among screened patients in a livestock dense area, 10.0% were MRSA carrier at a hospital test moment during the study period. Compared to the prevalence of MRSA carriage among general hospital patients at admission in different studies (0.11 – 0.15%) (Bode et al., 2011; Jurke et al., 2019; Wertheim et al., 2004; Workgroup Infection Prevention (WIP), 2012) this is at least 66 times higher. This suggests that risk groups included in the screening program are well-chosen, of which contact with livestock in particular, for this region. Four cluster areas were identified, where screened patients had a significantly (P<.05) higher MRSA carriage risk than those residing outside these cluster areas. The clusters were typically highly pig-dense.

Some discussion points have to be considered while interpreting the results, concerning the multicenter approach, geographical coverage and livestock densities.

An issue concerning the multicenter approach is the difference in hospital procedures regarding MRSA tests: some laboratories combined the throat and nose swab in one culture, while others cultured all swabs separately. Therefore, frequency of false negatives may differ per hospital, i.e. the number of carriers that are not recognized by testing. This may, to some extent, explain the heterogeneity in the hospital yields. Moreover, individuals that visit more than one of the included hospitals during the study period occur multiple times in our data. For future research it would be recommended for laboratories to harmonize their procedures more. A national initiative to have similar language in the laboratory information systems is already being implemented to facilitate such improvements. Recording risk factors that were reason to screen patients in the data system needs special attention.

Geographical clustering patterns could only be found among those residential four-digit postal codes in which at least 10 patients were screened (Appendix 4). The clusters A and B (Figure 4) were located within the province Gelderland; the other clusters were in Noord-Brabant where only one hospital in this region was included, such that some postal codes had less than 10 screened patients. Clusters tended to appear in non-urban areas. As screened patients had at least one of the risk factors for MRSA carriage and farmers usually live in rural areas, the clustering pattern might indicate that the risk for MRSA carriage is greater in (pig)farmers than in individuals with other risk factors, such as returning from a high-risk country. Coverage in Gelderland was rather good, with six out of nine hospitals included, but with a lack of coverage in the middle and North-East, such that possible clustering there remained unnoticed. While catchment areas of included hospitals largely overlap the catchment areas of the unincluded hospitals in central Gelderland, the relatively livestock-dense North-East is underrepresented, probably due to patients tending to visit hospitals in the province Overijssel instead.

Another point of discussion is that in the Province Gelderland, different types of livestock densities are spatially correlated, for example pigs and veal calve density, Pearson ρ = 0.37 (P< .001). Thus, although in the cluster A area there is a predominance of veal calve farming and in cluster C mostly pig farming, it is difficult to disentangle association of MRSA positivity with the different livestock densities as their farming areas overlap. In conclusion, we can state, as others previously have done, that living or working on a farm is a risk for MRSA carriage.

The fraction MRSA carriers among patients with livestock contact notified in the hospital system in this study was 22.7% and within the range of 9 to 30% mentioned in literature (Bureau Risicobeoordeling & onderzoeksprogrammering, 2012; IV et al., 2009; Mulders et al., 2010; van Cleef et al., 2010; Wassenberg et al., 2011; Workgroup Infection Prevention (WIP), 2012). Spread of livestock-MRSA to the community in livestock-dense areas in the Netherlands is not common, but related to the distance to a farm (Zomer et al., 2017). Our results suggest that the livestock denser an area is, the greater the MRSA carriage risk for the screened population at hospital admission.

Between 2009 and 2014, there was upscaling in pig farms, especially in Provinces Gelderland and Noord-Brabant: the number of farms declined, while the number of pigs did not diminish (Bosch et al., 2016). It would be interesting to investigate the role of pig farm size in MRSA carriage; if farm size does increase carriage risk, caution would be needed in upscaling of pig farms, to prevent the potential for MRSA carriage. This requires further analysis including time trends.

Of major concern is the bidirectional exchange between humans and pigs in farming environments because of its potential for new LA-MRSA strains (Price et al., 2012; Sieber et al., 2018). The common livestock-associated strain CC398 originated from humans as a non-resistant S. aureus, jumped to pigs and acquired resistance genes (Price et al., 2012). In recent years, Polish and Danish studies found MRSA strains other than CC398 in pigs, and in the United States in some states S. aureus strains other than CC398 make up the majority of typed isolates from pig farms (Mroczkowska et al., 2017; Sieber et al., 2018; Smith et al., 2018). Pig movements between breeding and production farms are found to be an important driver of MRSA spread and changing (MR)SA dynamics throughout pig farming industry in Denmark (Sieber et al., 2018). Pig movements within our study region possibly explain why clusters were found in some, but not in all pig-dense subareas. Usually LA-MRSA cases are considered independent introductions from the animal reservoir, where the spread is easier than in the human population (Donker et al., 2016). However, there is evidence for spread of a LA-MRSA subclade MT569 in the community, between persons without livestock contact in the Netherlands, living in four-digits postal codes that are not livestock-dense (Bosch et al., 2016).

## Conclusions

Among hospital-screened patients with a risk factor for MRSA carriage, the actual MRSA carriage risk is differentially distributed across the livestock-dense area within and surrounding Gelderland. Four MRSA carriage hotspots were identified in especially pig-dense areas, at the border of the province and to the south of the border. This implies that in this region, among livestock-associated risk factors having contact with pigs contributes most to the risk of MRSA carriage upon hospital admission. However, not all pig-dense areas in the hospital catchment areas were associated with a high-risk cluster. Research is needed into the role of pig movements between breeding and production farms, within the Netherlands and for export. Other future directions of work include the characterisation of regions with high livestock (pig) density but with low MRSA positivity, and examining the risk of MRSA carriage for the general population in the found hotspots. Potentially there are also lessons to be learned from farm dense areas with little MRSA transmission to humans. To disentangle the risks of contact with pigs, veal calves and broilers respectively, estimating the exposures at zip code level is not sufficient. Alternatives are to determine the individual exposure by the distance from the residence address to farms, a questionnaires or a GPS-tracker. Besides, it is advised that hospitals store the data on MRSA carriage risk factors and types of MRSA strains in a more systematic and harmonized way. This would greatly help in monitoring and adjusting the effectiveness of the MRSA screening policy in a low-cost and easy manner.

## Data Availability

The datasets generated and/or analyzed during the current study are not publicly available due to privacy protection of patients.

## List of abbreviations

MRSA: Methicillin resistant *Staphylococcus aureus*

## Declarations

### Funding

This study was funded by Radboudumc Nijmegen, and in kind by all the participating hospitals.

### Competing interests

The authors declare that they have no competing interests.

### Code availability

Code is available upon request.

### Ethics approval

The Radboudumc medical ethical committee approved the study.

### Consent to participate

Consent from included patients was not feasible. However, patients that raised objections against using their data for any later research by the time of testing, were not included in the original data.

### Consent for publication

Not applicable.

### Authors’ contributions

HW, HK, SF and EB initiated, supervised, reviewed and contributed with ideas to this project. TL, EM, MN, AV, AM, EJ, WS, MS and PS arranged agreement to obtain data from their hospitals and assisted with data collection on site. JH is responsible for MRSA screening in Radboudumc Nijmegen. VA further designed the project, monitored data collection, analyzed these and drafted this article.

## Acknowledgements

Hester Korthals Altes and Heiman Wertheim have contributed equally to this work. I would like to thank the following for their contributions: Ron Wunderink (GISdesk Radboud University Nijmegen) for the program ArcGIS; Pieter Timmerman (Department of Business Intelligence Radboudumc Nijmegen) for catchment area data that were used in a pilot study; Thijs Bosch (RIVM) for a list of LA-MRSA associated Spa-types; Twan Klaassen, Jeanette Rijksen, Willeke Schellekens, Jamie Berendsen, Monique Bruns, Maarten van Mourik, Marcel Tonissen, Agi Marik, Patricia Buitels, Sandra Thonen, Thea Teijgeler, Jaap from Slingeland Doetinchem, Nelleke van der Weerd (included hospitals) for their efforts for committee approvals and data processing.

## Notes

### Competing Interest Statement

The authors have declared no competing interest.

### Author Declarations

The Radboudumc medical ethical committee approved the study.

