## Supplements for "Spatial analysis of methicillin-resistant *Staphylococcus aureus* carriage (MRSA) at hospital admission in a livestock dense region"

### Appendix 1 Notes on coverage

Initially, nine hospitals participated in this study. Data storage differed between hospitals, as a result of different lab systems and test procedures. Moreover, lab systems are built to facilitate viewing individual patient information rather than providing large data files. For two hospitals, we were unable to retrieve data in a manner comparable with the remaining hospitals. Therefore, the authors decided to exclude these from the main analysis. Figure A1 visualizes the overlap in treatment areas between the hospitals A-G (included, Fig. A1a-g), and H-I (excluded, Fig. A1b and Fig. A1c respectively). It is apparent that patients screened in hospital I concentrate in a central belt in Gelderland, while hospital H screens with lower intensity over scattered areas in Gelderland and Noord-Brabant.

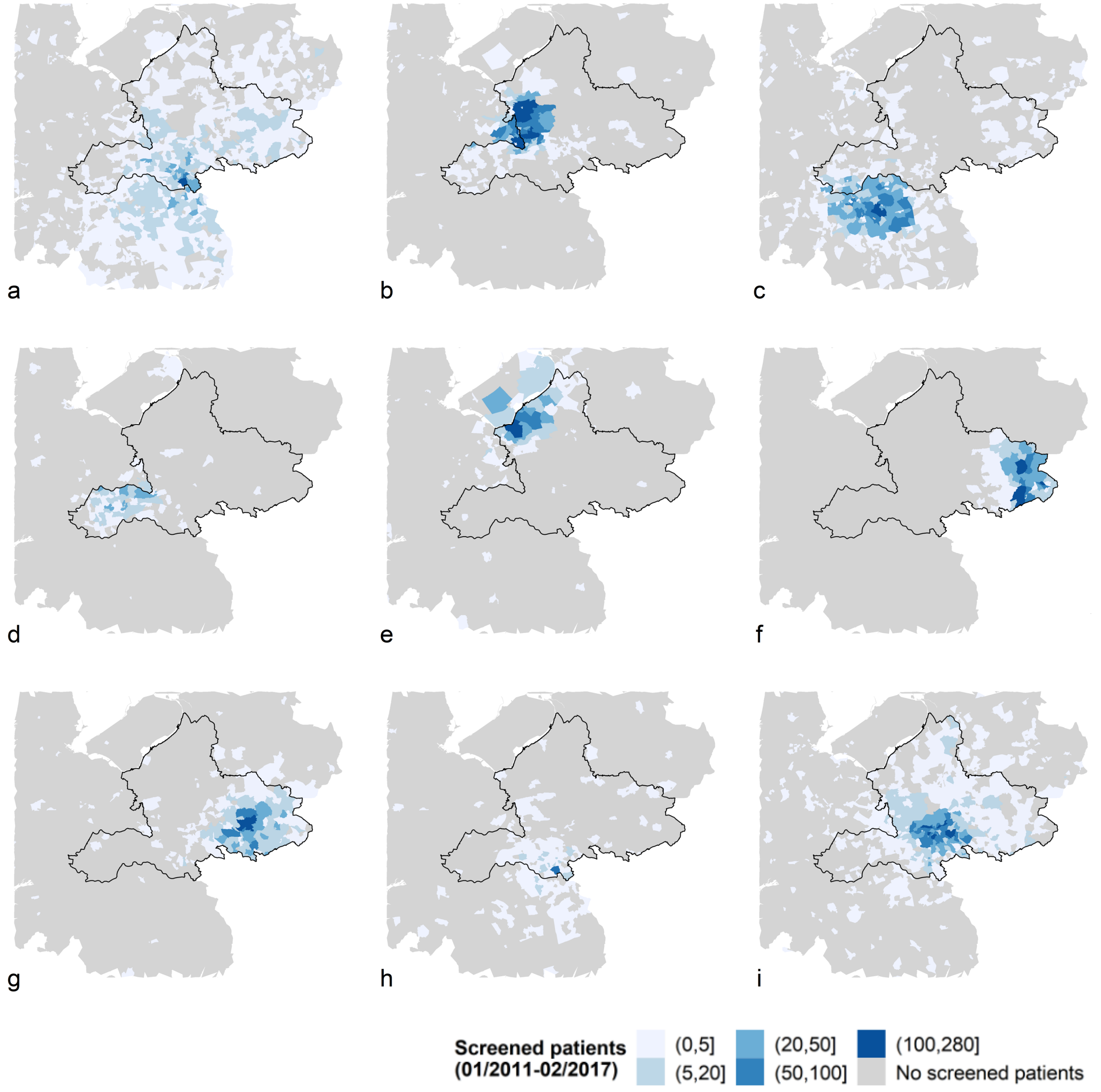


**Figure A1** MRSA-screened patients per 4 digit postal code area of nine hospitals in the period 01/2011-02/2017 . a-g) hospital A to G (included); h-i) hospital H and I (excluded). four-digits postal codes in which at least 10 patients were screened, were included. The hospital letters do not correspond to those in Table AI to suffice anonymity. The maximum number of screened patients per postal code per hospital is 280.

### Appendix 2 Age and sex

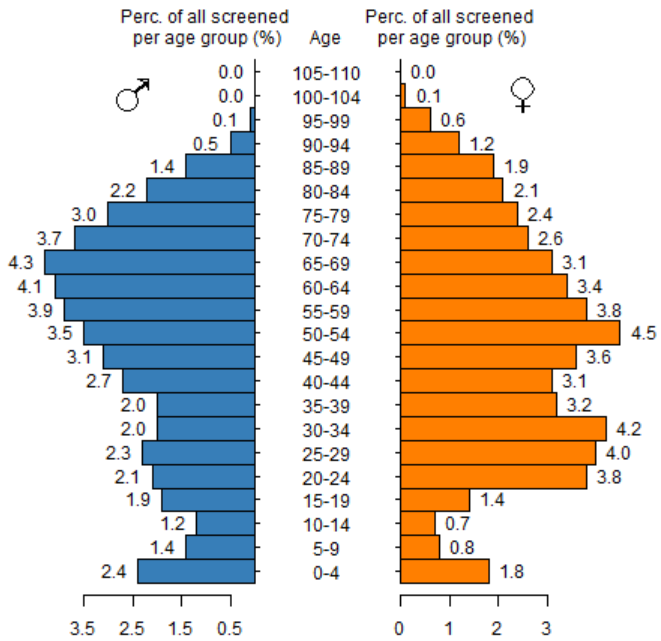


**Figure A2** Age distribution of patients (% of total), screened for MRSA carriage. Percentages are calculated over the total count of females and males. For 82% (n=17 565) of included patients, age and sex were known.

### Appendix 3 SaTScan spatial scan

Before running the model, three datasets were specified with the R package 'rsatscan' (22). The cases file consisted of the count of MRSA carriers per age/sex group per four-digits postal code; the geographical file consisted of the X- and Y-coordinates of the centroids of all four-digits postal codes where at least one patient was screened, such that only these four-digits postal codes would be taken along by the scanning window; the population file consisted of the count of screened patients per age/sex group per four-digits postal code. The spatial scan statistic identifies clusters by centering a circular window of flexible radius in the centroid of the four-digits postal code areas, and comparing expected and observed numbers of cases inside and outside the window. We used the Poisson model which calculates the expected cases by assuming that the number of cases in each area is Poisson distributed. P-values were calculated by performing 999 Monte Carlo simulations (default option) and clusters with $P<.05$ were selected. Secondary clusters were only reported if there was no geographical overlap with the most likely cluster. The maximum percentage of population at risk in a cluster was 10%, and the maximum radius per cluster was 10 km. The detected clusters were superimposed on the MRSA carriage incidence map as circles. Moreover, livestock density maps were drawn and clusters were superimposed on these maps in the same way to examine the livestock densities in four-digits postal codes of MRSA carriage hotspots and outside of these.

### Appendix 4 Summary per hospital

**Table AI** Summary findings per hospital (anonymous). The total number of screened patients with admission/inpatient visit in the period of 01/2011-02/2017, the number of patients for which the risk factor that was reason for screening was given in the data and the number of found MRSA carriers as part of the screening program are shown per included hospital. Percentages are calculated with screened patients as denominator. Hospitals A and I (letters are different from additional file 1) were excluded for analysis.

| Hospital | Screened patients | Risk factor known | (% of screened pat.) | MRSA carriers | (% of screened pat.) |
|---|---|---|---|---|---|
| A | 5 476 | 1 211 | 22.1 | 33 | 0.6 |
| B | 3 848 | 0 | 0 | 426 | 11.1 |
| C | 3 411 | 133 | 3.9 | 314 | 9.2 |
| D | 3 255 | 97 | 3.0 | 277 | 8.5 |
| E | 1 973 | 679 | 34.4 | 113 | 5.7 |
| F | 1 336 | 27 | 2.0 | 188 | 14.1 |
| G | 1 050 | 0 | 0 | 125 | 11.9 |
| H | 673 | 30 | 4.5 | 56 | 8.3 |
| I | 391 | 208 | 53.2 | 161 | 41.2 |
| Total | 21 413 | 2 385 | 11.1 | 1 693 | 7.9 |

Cluster D found with the larger set of hospitals does coincide with a pig-dense area (Fig.A3b and Fig. 4b) not covered by a cluster identified with the limited subset of hospitals (fig, 4b). We also observe a reduction in the diameter of the cluster C in fig, A3b, which corresponds to the area of cluster B in the subset analysis (fig., 4b). Strangely enough, this is difficult to understand, because Fig. A1b and c does not reveal many screened cases in the area of cluster C. Clusters F and G from the analysis with all hospitals correspond to pig-dense areas. The area with many screened patients for the excluded hospital indicated in fig. A1b (a belt-like area in Gelderland), does not correspond with a pig-dense area (or live-stock dense; fig. 4b-c) and is not associated with a cluster of higher MRSA-positivity.

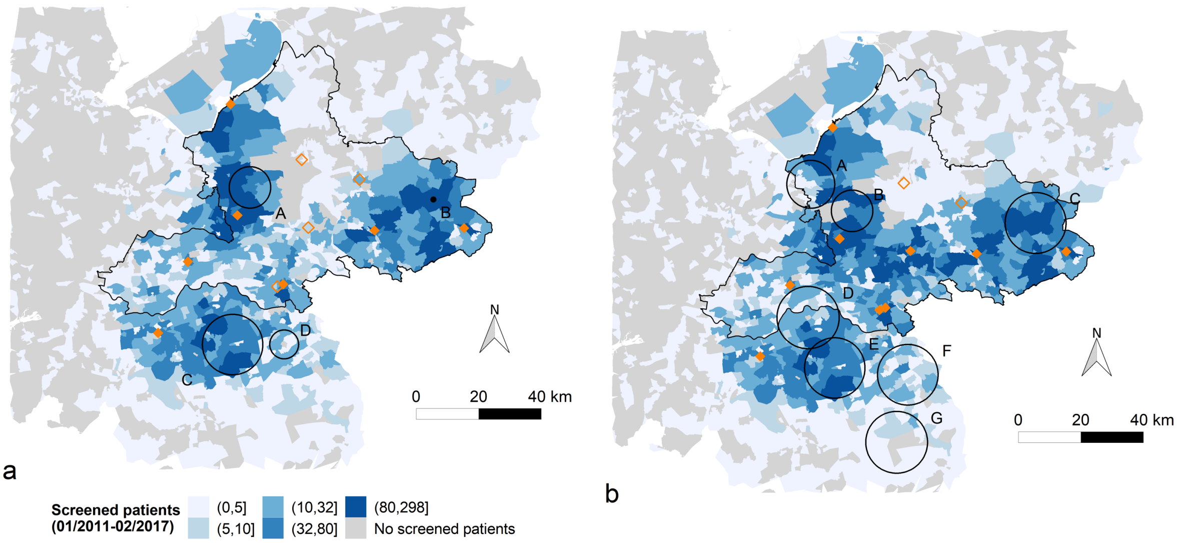


**Figure A3** MRSA screened patients per postal code and clustering in the period 01/2011-02/2017. Clusters of MRSA carriage are superimposed on maps with the absolute number of patients screened per four-digits postal code based on the respective data set: (a) Seven hospitals (main analysis); (b) Nine hospitals (initially included). SaTScan analysis was performed for four-digits postal codes with at least 10 screened patients only. Orange (filled) diamonds represent included hospitals, open diamonds represent hospitals excluded for analysis. The border of the Province Gelderland is indicated in black.
